# Classical driver mutations are not associated with metachronous lesion risk in patients undergoing post-polypectomy surveillance following removal of conventional adenomas in a bowel screening setting

**DOI:** 10.64898/2026.08.05.26359768

**Authors:** Stephen T McSorley, Leonor P Schubert Santana, Aula Ammar, Sara S.F. Al-Badran, Emma C Parsons, Philip D Dunne, Noori Maka, Mark Johnstone, Gerard Lynch, Joanne Edwards

**Author notes:** Corresponding author: Dr Stephen T McSorley, Senior Clinical Research Fellow and Honorary Consultant Colorectal Surgeon School of Cancer Sciences, University of Glasgow, Wolfson Wohl Cancer Research Centre Bearsden, Glasgow G61 1BD, UK.

## Abstract

**Introduction:** Patients undergoing polypectomy at colonoscopy remain at risk of metachronous neoplasia despite surveillance guided by histopathological features. Mutational profiling of adenomas, including canonical driver mutations in *APC, KRAS*, and *TP53*, may offer additional predictive value. This study aimed to determine whether mutational status in index adenomas was associated with metachronous lesion risk.

**Methods:** The INCISE cohort included patients aged 50–74 who underwent polypectomy within the Scottish Bowel Screening Programme and subsequent surveillance colonoscopy within 6 years. Targeted next-generation sequencing was performed on formalin-fixed paraffin-embedded polyps. Driver mutation frequency, tumour mutational burden (TMB), and variant allele frequency (VAF) were analysed and correlated with histopathological features and metachronous outcomes using appropriate statistical models.

**Results:** A total of 895 adenomas from 723 patients were analysed. In conventional adenomas, as the number of high-risk histopathological features (size ≥10mm, villous architecture, and high-grade dysplasia) increased there was a stepwise increase in the proportion of samples with a mutation in *KRAS* from 13% to 51% (padj<0.001) and *TP53* from 8% to 35% (padj<0.001). However, neither mutation frequency (p=0.901), nor median tumour mutation burden (TMB) (2.27 vs 2.15 mut/Mb, p=0.242), in index adenomas was associated with the development of metachronous lesions.

**Conclusions:** While classical driver mutations reflect histopathological progression within adenomas, they do not predict metachronous lesion risk post-polypectomy. Targeted mutation profiling alone is insufficient for surveillance risk stratification, highlighting the need for integrated molecular approaches in this setting.

## Introduction

Colorectal cancer (CRC) is among the most prevalent malignancies worldwide [1]. Evidence suggests that population-based screening programmes have reduced cancer specific mortality through early detection, and have reduced CRC incidence in screening age patients through the removal of pre-cancer polyps [2–3]. However, patients who have undergone polypectomy remain at risk of future colonic neoplasia [4], necessitating surveillance in those considered to be at high-risk [5], to improve long term outcomes [6].

Current post-polypectomy surveillance guidelines internationally are founded on histopathological features of the largest most advanced index lesion - principally size, degree of dysplasia, villous architecture - and number of synchronous adenomas [7]. However, they provide incomplete risk stratification, with a significant proportion of patients classified as low-risk developing metachronous advanced adenomas or interval cancers, whilst many classified as high-risk do not and so undergo unnecessary surveillance [8, 9].

The **IN**tegrated **T**e**C**hnologies for **I**mproved Polyp **S**urveillanc**E (**INCISE) patient cohort, identified from the Scottish Bowel Screening Programme in NHS Greater Glasgow and Clyde, was established to determine whether molecular, immunological, and digital pathology features of index adenoma tissue could improve on histopathological risk stratification for metachronous neoplasia [9]. A systematic review of molecular and non-histopathological markers of metachronous lesion risk published at the outset of the project identified several candidate biomarkers, including key driver mutations associated with the adenoma carcinoma sequence, *APC*, *KRAS* and *TP53* [10].

The most common molecular subtype of CRC (chromosomal instability, CIN, pathway) is characterised by a lengthy premalignant phase, during which most sporadic tumours arise through the progressive transformation of conventional adenomatous polyps, providing the interception window for the aforementioned screening and surveillance strategies [11, 12]. This process was originally described by Fearon and Vogelstein as the adenoma-carcinoma sequence (ACS), a stepwise model in which the accumulation of somatic driver mutations in *APC, KRAS, SMAD4*, and *TP53* drives sequential histopathological and biological progression from normal epithelium through adenoma to invasive carcinoma [13]. Whilst this model has been enormously influential, it is now recognised to be an oversimplification.

Multi-region sequencing and single-cell studies have demonstrated heterogeneity within and between adenomas, with driver mutations being sub-clonal rather than clonally fixed.

Progression to carcinoma is driven mainly by chromosomal changes rather than point-mutations, with additional modulation from the tumour microenvironment and the surrounding mutagenic field of the mucosa [14–16]. Collectively this means that the relationship between somatic driver mutation status in an individual adenoma and that adenoma’s biological trajectory is substantially more complex than the canonical ACS implies [17–19].

Targeted sequencing of polyps longitudinally has revealed that small adenomas accruing driver mutations at baseline are associated with subsequent growth and high-grade change, suggesting that mutational profiling may have prognostic value in specific contexts [20].

Integrated genomic sequencing as part of the GENESIS study demonstrated that a genetic biopsy approach incorporating mutational and methylation data could predict surveillance intervals after polypectomy [21]. However, it remains unclear whether overall mutational burden, frequency of specific mutations, or sequential accrual of canonical ACS driver mutations in index adenomas independently predict metachronous lesion risk after accounting for histopathological features in a large, well characterised screening cohort.

The present study addresses these questions in patients enrolled in the INCISE cohort. We characterised driver mutation profiles in index, synchronous, and metachronous conventional adenomas using a targeted next-generation sequencing panel and examined the associations of these molecular features with advanced histopathological characteristics and with the detection of metachronous neoplasia during surveillance.

## Methods

### Patient cohort

The INCISE cohort includes patients aged 50-74 years, who between 2009 and 2016 underwent polypectomy within NHS Greater Glasgow and Clyde as part of the biennial Faecal Immunochemical Test (FIT) based Scottish Bowel Screening Programme, and had at least one surveillance colonoscopy between 6 months and 6 years later. Patients were identified and data linked using the unique Community Health Index (CHI) number. Patient demographic and clinical information were recorded from the electronic health record (Clinical Portal, Orion Health). Data including polyp number, morphology, size, location and surveillance outcome were recorded from electronic endoscopy reporting software (Unisoft GI Reporting Software, v2.5, Unisoft Medical Systems). Polyp histology and dysplasia were collected from the electronic pathology database (Telepath Laboratory Information Management System). Patients with colorectal cancer (CRC), history of CRC, a known polyposis or CRC risk syndrome or inflammatory bowel disease (IBD) were not included.

### Tissue processing and DNA sequencing

The most advanced polyp removed at the first bowel screening colonoscopy was identified on the basis of lesion size and highest grade of dysplasia, and the corresponding Formalin Fixed and Paraffin Embedded (FFPE) samples retrieved from the clinical pathology archive and identified as “index”. Other polyps from the same procedure were identified as “synchronous” and were similarly retrieved. Finally, FFPE samples of polyps removed at the surveillance procedure were also retrieve and identified as “metachronous”.

Haematoxylin and Eosin (H&E) FFPE whole slide adenoma sections were scanned at 40x resolution at the Glasgow Tissue Research Facility (GTRF) with a Hamamatsu S60 NanoZoomer (Hamamatsu, Japan) and images were stored within NZConnect (v 1.1.0) and accessed using NDP Viewer.

DNA was extracted from FFPE polyp samples using a Maxwell CSC FFPE (Promega) with a minimum acceptable DNA concentration of 1.0 ng µl^−1^ determined by Qubit 4 Fluorometer, (Thermo Fisher Scientific). Sequencing and variant calling were performed by the Genomic Innovation Alliance (GIA) using the Agilent SureSelect XT2 HS2 method (Agilent). Regions of interest were enriched with the Agilent SureSelect CancerPlus panel (design ID: S3225252) and the quality and quantity of libraries was determined using a D1000 ScreenTape on TapeStation (5067–5582, Agilent).

Sequencing data were processed and SNV files were generated using GIA’s proprietary HOLMES pipeline (v.1.3.1). Bcl2fastq conversion software (Illumina, v.2.19.1.403, v.2.20.0.422 on C++) was used to convert NovaSeq-6000-generated .cblc raw data files to FASTQ files, which were then aligned using Burrows–Wheeler Alignment (v.0.7.15 as a C programme). deepSNV/Shearwater R Package/R wrapper script (v.1.22/v.1.1.0, v.1.22.0.5/v.1.1.0) was used for calling SNVs, while Pindel (v.0.2.5b8-ww1 as a standalone application) was used for large insertions/deletions. Variants were then annotated using CAVA (v.1.2.2.ww1, v.1.2.2.ww5 on Python). VCF files were converted to MAF format using vcf2maf (v.1.6.22), and VEP (v.112) was used for reannotation.

Mutations were annotated as drivers if they were not a common variant, were reported in 3 or more samples, and were called either “oncogenic” or “likely oncogenic” in COSMIC [22] or OnkoKB [23], excluding variants in HLA genes. Variant allele frequency (VAF) was estimated as the proportion of reads supporting the alternate allele of all reads at that location, without correction for adenoma cellularity and without dedicated copy number data given the limitations of the targeted panel used.

### Statistical analysis

Somatic mutation data were analysed in R (v4.5.2) using the maftools R package (v2.26.0)[24] under the BiocManager Repository (Ver. 1.30.22)[25]. Samples were included if they met quality control criteria of average coding depth >50x.

Mutation landscape analyses included oncoplot summaries generated using maftools with only variants annotated as driver events retained. Stacked bar comparisons were performed between clinical subgroups using coBarplot function within maftools, with Fisher’s exact test or McNemar’s test applied for unpaired and paired comparisons respectively, with Benjamini-Hochberg (BH) false discovery rate correction.

Oncogenic pathway analyses were performed using maftools (v2.26.0) by computing the proportion of samples harbouring at least one mutation in curated gene sets for ten canonical cancer pathways (WNT, RTK/RAS, TP53, PI3K, Cell Cycle, MYC, Hippo, Notch, Splicing, TGF-beta)[26].

Tumour mutational burden (TMB) was calculated using the tmb function with a panel capture size of 3.96Mb, counting non-silent mutations and excluding drivers. TMB was visualised using violin plots created by ggplot2 (v4.0.1)[27] and compared across clinical subgroups including timepoint (index, synchronous, metachronous), future polyp or colorectal cancer (CRC) detection, dysplasia grade, polyp size, and histological subtype using Wilcoxon signed rank test for pairwise comparisons, Wilcoxon rank-sum test for unpaired two group and Kruskal-Wallis test for more than two group comparisons.

VAF estimate distributions were compared at the gene level and visualised using density plots described by easyVAF [28], with the value from the locus with maximum VAF estimate used in samples with multiple mutations in the same gene.

Venn diagrams of sample timepoint and advanced feature overlap were created using ggVennDiagram (v1.5.7)[29]. An advanced histological feature score (0-3) was constructed by summing binary indicators for the presence of: polyp size ≥10mm, tubulovillous or villous histology, and high-grade dysplasia. The association between mutation frequency of the top 10 most frequently mutated genes and this ordinal score was assessed per gene using Cochran-Armitage trend tests with Benjamini-Hochberg correction, and TMB compared using the Kruskal Wallis test.

Paired analyses comparing index to synchronous adenomas and index to metachronous adenomas were performed first using patient-matched samples. TMB comparisons in matched pairs used the Wilcoxon signed-rank test. Paired cobar plot analyses used McNemar’s test with Benjamini Hochberg correction. To account for potential confounding by clinical covariates, two independent propensity score matched analyses were performed using MatchIt [30], comparing index to synchronous adenomas; and index to metachronous adenomas. Propensity scores were estimated for each pairwise comparison using logistic regression with the following covariates: sex, colonic anatomical location, histological subtype, size, and dysplasia grade. For each analysis, synchronous or metachronous samples were individually matched to their nearest available index sample using nearest-neighbour matching without replacement. An absolute caliper of 0.01 on the propensity score (0–1) scale was applied. Post-matching covariate balance was assessed by computing the absolute standardised mean difference (ASMD) for each covariate before and after matching. Within the matched samples, TMB was compared using the Wilcoxon signed-rank test, and mutation frequency was compared between groups using McNemar’s test with Benjamini-Hochberg correction.

Finally, index adenomas were grouped by three classic adenoma-carcinoma sequence driver mutations into those without an *APC* mutation (No APC), at least one *APC* mutation (A), at least one *APC* and one *KRAS* mutation (AK), and at least one *APC* and one *KRAS* and one *TP53* mutation (AKP). Kaplan-Meier analysis of time to future polyp or CRC was performed using SPSS (v29, IBM, Armonk), with groups compared using the log rank test.

All statistical tests were two-sided; p<0.05 was considered statistically significant.

### Approvals

Ethical approval was granted by the West of Scotland Research Ethics Committee and NHS Greater Glasgow and Clyde SafeHaven for the use of surplus diagnostic tissue (22/WS/0020) and retrospective data (GSH/20/CO/002) without individual level informed consent respectively.

## Results

### Patient and sample cohort

DNA was extracted and gene panel mutation data generated for a total of 1033 polyps from 791 patients (Figure 1). 101 samples from 45 patients were excluded due to having an average sample read depth of <50x, and a further 37 samples from 23 patients which were not conventional adenomas were excluded. This gave a final total of 895 adenomas from 723 patients, of which 701 were index, 148 were synchronous and 46 were metachronous (Table 1). Of the 723 patients, 534 (74%) were male and 189 (26%) were females, with a median age of 63 yrs (IQR 57-69) at index bowel screening colonoscopy, and of which 434 (60%) were found to have a metachronous polyp or CRC during surveillance. Mutations in genes associated with the *Wnt* signalling pathway (92.7% of adenoma samples) were most frequent, followed by mutations in genes associated with *RTK/RAS* (41.5%) and *TP53* (22.4%) pathways (Supplementary Figure 1). There were significant differences in adenoma location, and high-risk features including size ≥10mm, villous histology (TV/V), high grade dysplasia (HGD) between timepoints (all p<0.001), with the greatest differences found in synchronous samples which had lower frequency of high-risk features when compared to either index or metachronous (Table 1).

**Figure 1:**
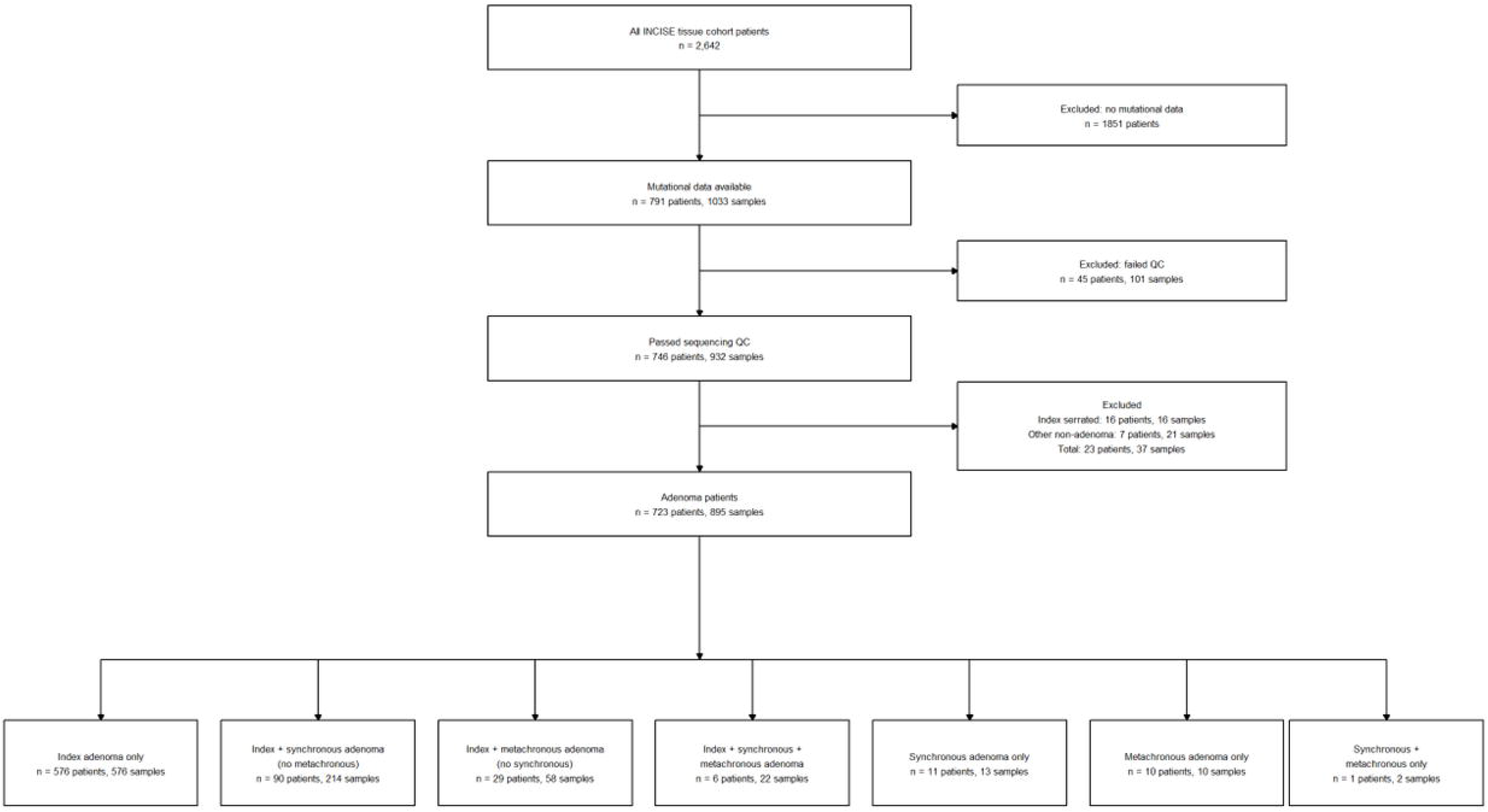
Patient and sample inclusion flow chart of INCISE mutational data cohort.

**Table 1:** Conventional adenoma characteristics by sampling timepoint from INCISE tissue cohort with available mutational data.

| Time point |  | Index | Synchronous | Metachronous | p* |
| --- | --- | --- | --- | --- | --- |
| **Samples (n)** | - | 701 | 148 | 46 | - |
| **Size n(%)** | <10mm | 56 (8) | 64 (43) | 4 (9) | <0.001 |
|  | ≥10mm | 645 (92) | 84 (57) | 42 (91) |  |
| **Location n(%)** | Right | 119 (17) | 48 (32) | 14 (31) | <0.001 |
|  | Left | 501 (71) | 75 (51) | 23 (50) |  |
|  | Rectum | 78 (11) | 22 (15) | 7 (15) |  |
|  | Unknown | 3 (1) | 3 (2) | 2 (4) |  |
| **Histology n(%)** | T | 302 (43) | 110 (74) | 17 (37) | <0.001 |
|  | TV | 356 (51) | 35 (24) | 25 (54) |  |
|  | V | 43 (6) | 3 (2) | 4 (9) |  |
| **Dysplasia n(%)** | LGD | 595 (85) | 148 (100) | 40 (87) | <0.001 |
|  | HGD | 106 (15) | 0 (0) | 6 (13) |  |

HGD high grade dysplasia, LGD low grade dysplasia, T tubular, TV tubulovillous, V villous
\*chi square

### Advanced histopathological features of index conventional adenomas are associated with mutations in classic drivers KRAS and TP53

Overlap of the three high-risk characteristics, size ≥10mm, tubulovillous or villous histology, and high-grade dysplasia (Figure 2A) was common with only 10% (n=87) of adenoma samples having no high-risk features and of those with any high-risk feature, 57% having 2 or more (n=467)(Figure 2B). When samples were assigned a score from 0 to 3 based on the number of high-risk characteristics present, as the number of high-risk characteristics increased there was a stepwise increase in the proportion of samples with a mutation in *KRAS* from 13% to 51% (padj<0.001) and *TP53* from 8% to 35% (padj<0.001) (Figure 2C). There was no significant difference in TMB (Figure 2D, p=0.094).

**Figure 2:**
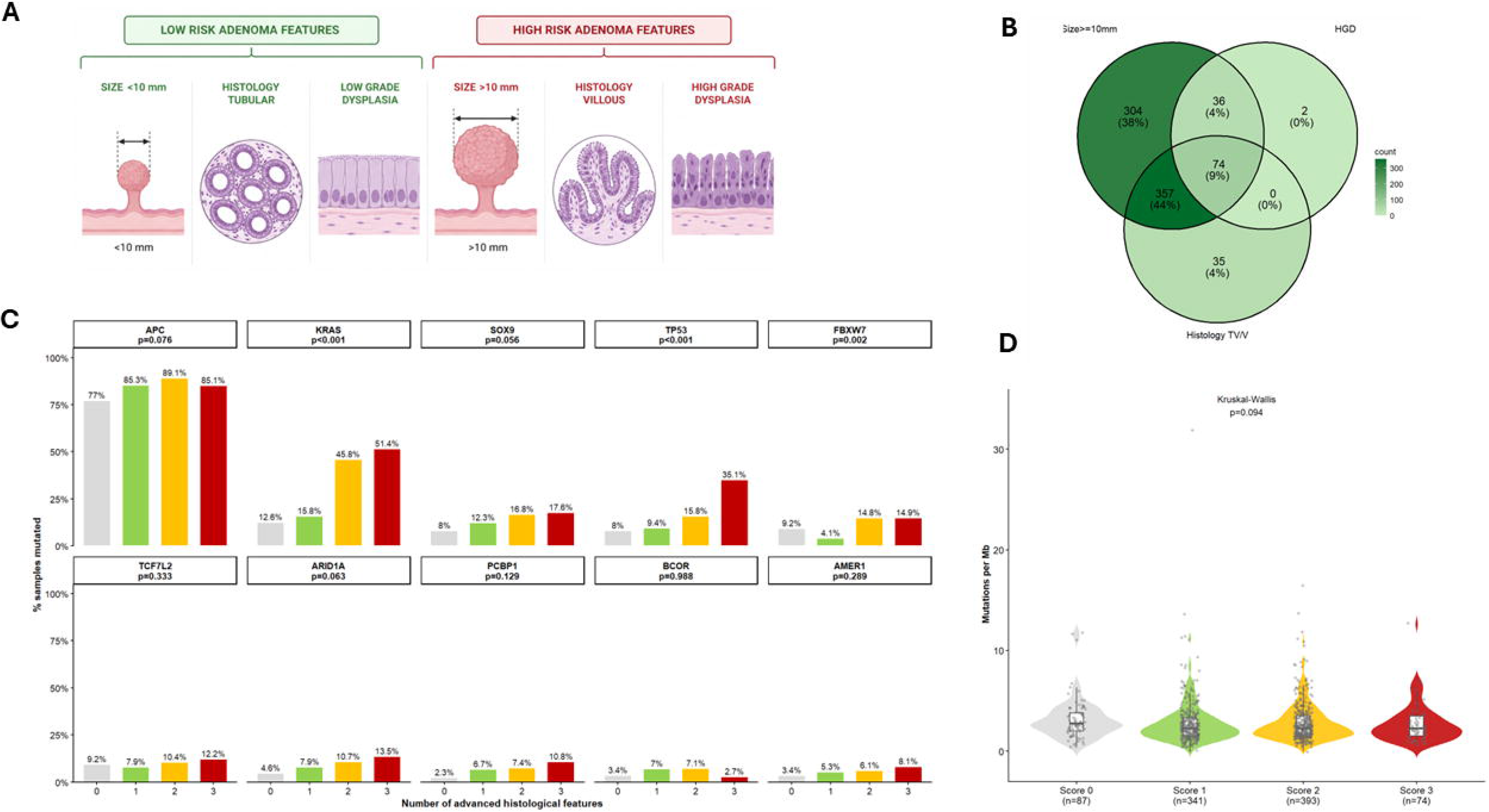
Mutation frequency is associated with histological features of advanced lesions in conventional adenomas. **(A)** Schematic of high risk histological features of conventional adenomas. **(B)** Venn diagram of histopathological features associated with advanced conventional adenomas; size ≥10mm, high grade dysplasia (HGD) and villous featured (TV/V). **(C)** Histograms comparing frequency of a mutation in each of the top 10 mutated genes (panels) between samples with 0 (grey), 1 (green), 2 (yellow) or all 3 (red) advanced features shows significantly higher frequency of mutations in *KRAS* (padj<0.001), *TP53* (padj<0.001) and *FBXW7* (padj=0.002) as the number of advanced features in a sample increases (Cochran Armitage test for trend with Benjamini-Hochberg adjustment). **(D)** Violin plot comparing distribution and medians of tumour mutational burden (TMB) between samples with 0 (grey, n=87, 2.78 mut/Mb), 1 (green, n=341, 2.27 mut/Mb), 2 (yellow, n=393, 2.27 mut/Mb) or all 3 (red, n=74, 2.27 mut/Mb) histological features of advanced lesions, based on a panel size of 3.96Mb (Kruskal-Wallis p=0.094).

### Mutation burden but not driver mutation frequency vary between index, synchronous and metachronous adenomas

Index, synchronous and metachronous adenomas were compared (Figure 3A). Per patient matching resulted in n=96 matched index to synchronous and n=35 matched index to metachronous pairs (Figure 3B). There were significant differences in TMB between index (median 2.53 mut/Mb) and synchronous (median 3.03 mut/Mb) adenomas from the same patient (p=0.003, Figure 3C), and between paired index (median 2.02 mut/Mb) and metachronous (median 3.79 mut/Mb) adenomas from the same patient (p=0.003, Figure 3D). There were no significant differences in the frequency of the top 20 most mutated driver genes when paired index and synchronous (Figure 3E) or index and metachronous (Figure 3F) adenomas from the same patient were compared.

**Figure 3:**
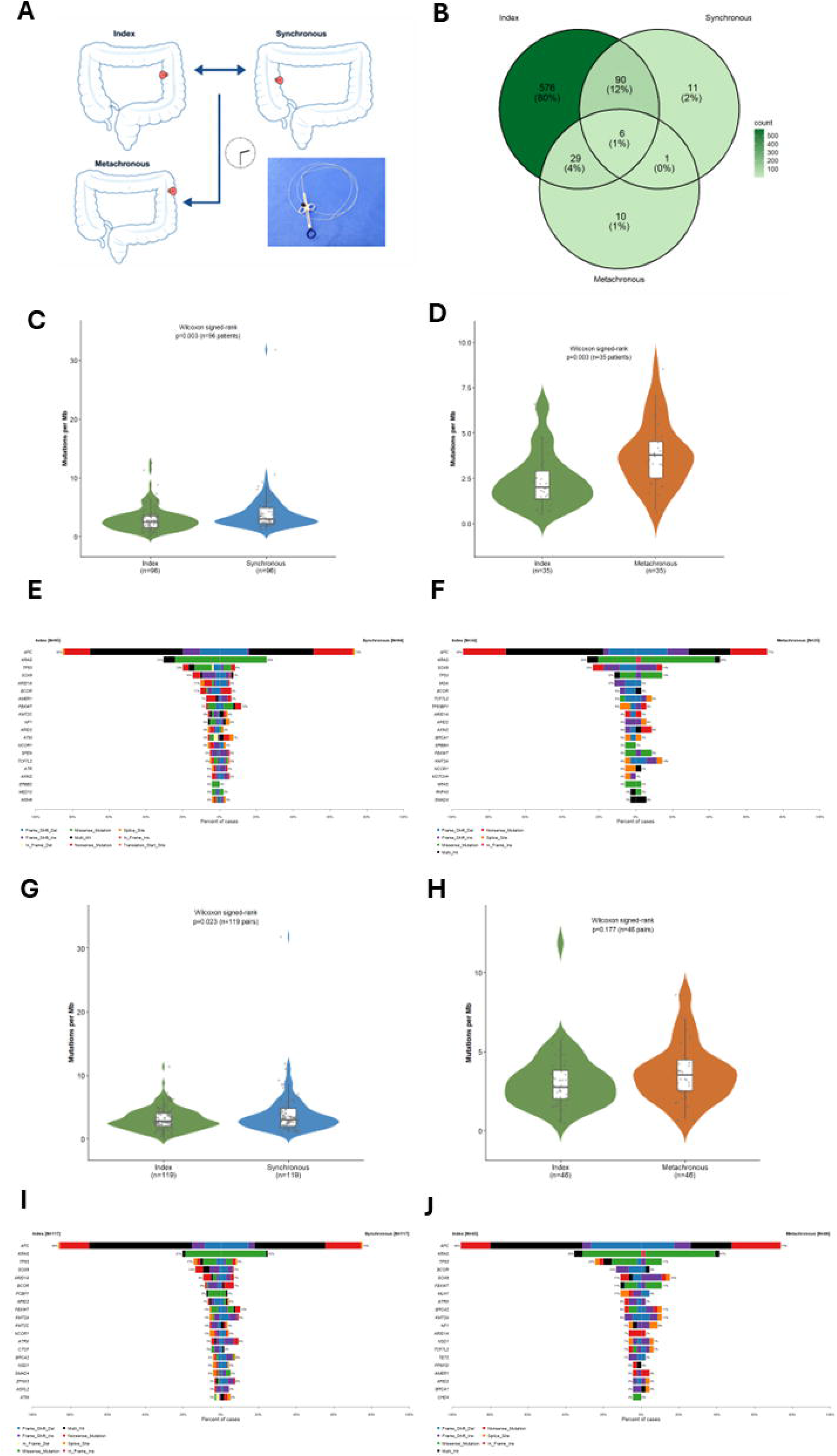
Matched comparison of mutations across index, synchronous and metachronous conventional adenomas. **(A)** Schematic of space and time point sampling of index, synchronous and metachronous adenomas. **(B)** Venn diagram of overlap of conventional adenoma sampling per patient. **(C)** Violin plot comparing distribution and median tumour mutational burden (TMB) between per- patient paired index (median 2.53 mut/Mb) and synchronous (median 3.03 mut/Mb) adenomas from the same patient (Wilcoxon signed rank p=0.003) **(D)** Violin plot comparing distribution and median tumour mutational burden (TMB) between per-patient paired index (median 2.02 mut/Mb) and metachronous (median 3.79 mut/Mb) adenomas from the same patient (Wilcoxon signed rank p=0.003). Cobar plots of top 20 mutated genes comparing **(E)** per-patient paired index and synchronous (n=96) conventional adenomas and **(F)** per-patient paired index and metachronous adenomas (n=35) showing no significant difference in mutation frequency (McNemar’s test with Benjamini-Hochberg adjustment). **(G)** Propensity score matched violin plot comparing distribution and median tumour TMB between paired (n=119) index (median 2.78 mut/Mb) and synchronous (median 3.03 mut/Mb) adenomas (Wilcoxon signed rank p=0.023) **(H)** Propensity score matched violin plot comparing distribution and median tumour TMB between paired (n=46) index (median 2.78 mut/Mb) and metachronous (median 3.54 mut/Mb) adenomas (Wilcoxon signed rank p=0.177). Cobar plots of top 20 mutated genes comparing **(I)** histological feature propensity score matched index and synchronous adenomas, and **(J)** histological feature propensity score matched paired index and metachronous adenomas with no significant differences in frequency (McNemar’s test with Benjamini Hochberg correction).

As an alternative to per patient matching, a per polyp matching was carried out using propensity scoring based on clinical and pathological characteristics for index and synchronous (n=119 pairs) and index and metachronous (n=46 pairs) adenomas, regardless of patient of origin, resulting in significant improvements in covariate balance for both (Supplementary Figure 2). Propensity score matched comparison of TMB between paired index (median 2.78 mut/Mb) and synchronous (median 3.03 mut/Mb) adenomas (Figure 3G), showed a significant difference (p=0.023). In contrast, propensity score matched comparison of median tumour TMB between paired index (median 2.78 mut/Mb) and metachronous (median 3.54 mut/Mb) adenomas (Figure 3H) found no significant difference (p=0.177). Propensity score matched cobar plots of top 20 most frequent driver mutations comparing paired index and synchronous adenomas (Figure 3I), and paired index and metachronous adenomas (Figure 3J) showed no significant differences in mutation frequency.

### Mutation frequency or burden in index conventional adenomas are not associated with likelihood of detection of metachronous lesion during surveillance

Considering only the index adenoma per patient, there was no significant difference (p=0.242) in TMB (Figure 4A), or in the frequency of the top 20 most commonly mutated genes (Figure 4B) when those who did and did not have a future polyp or CRC detected during surveillance were compared. At multivariate time to event analysis (Figure 4C), male sex (HR 1.38, 95% CI 1.11-1.72), and both left colon (HR 0.60, 95% CI 0.48-0.75) and rectal (HR 0.59, 95% CI 0.42-0.82) location with reference to right colon were significantly associated with time to detection of metachronous lesion during surveillance, whilst none of the three high risk histological characteristics were associated with this outcome.

**Figure 4:**
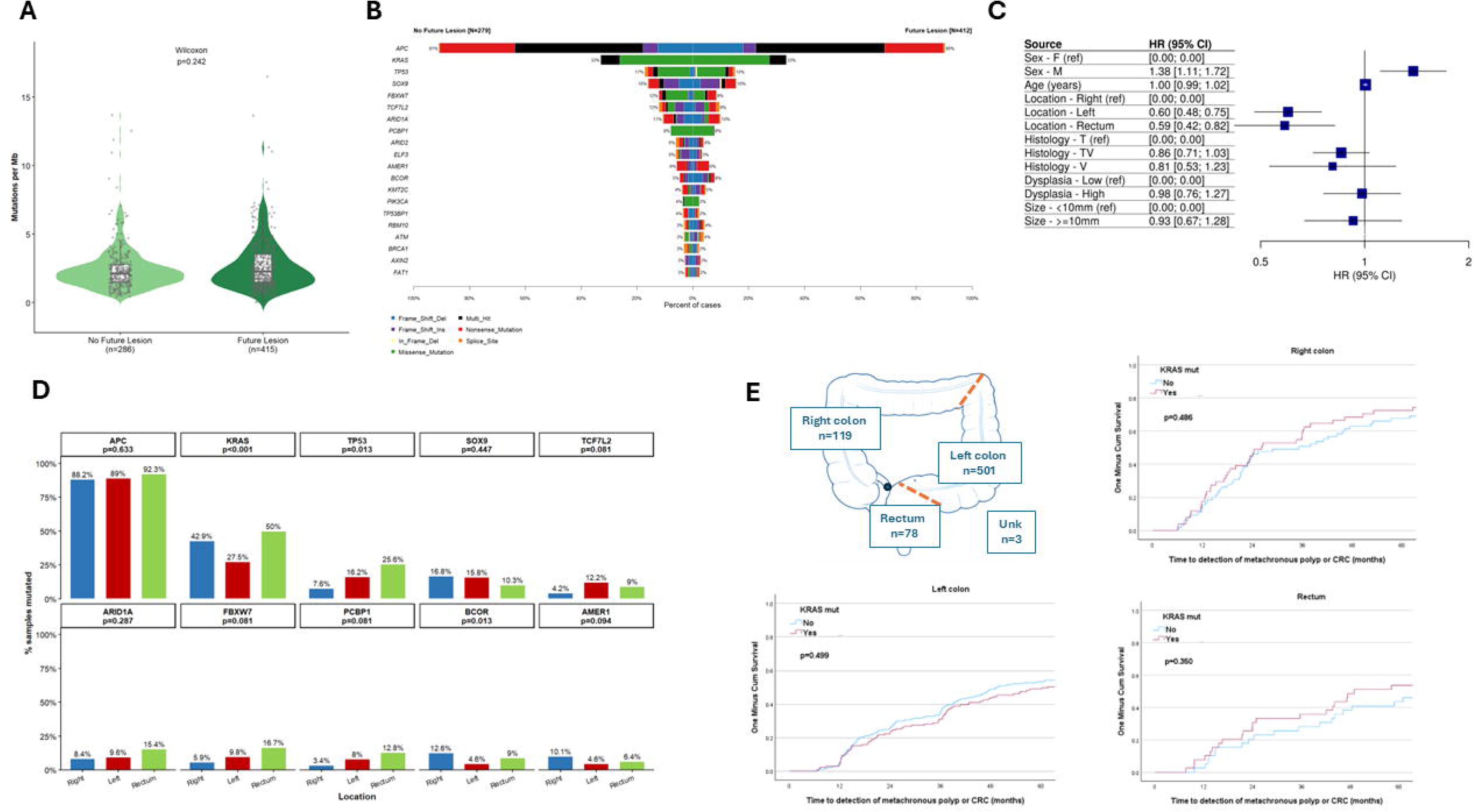
No difference in mutations within index conventional adenomas in relation to likelihood of metachronous polyp or colorectal cancer. **(A)** Violin plot of distribution of tumour mutational burden (TMB), showing no association with future polyp or CRC (median 2.27 vs 2.15 mut/Mb), based on a panel size of 3.96Mb, Wilcoxon rank-sum p=0.242. **(B)** Cobar plot of top 20 mutated genes in index conventional adenomas showing no significant difference in mutation frequency between no future polyp or CRC and future polyp or CRC (Fisher’s exact test with Benjamini Hochberg adjustment). **(C)** Forest plot displaying Multivariate Cox regression hazard ratios (HR) and 95% confidence intervals (CI) of patient and adenoma characteristics associated with future polyp or CRC shows significant association with male sex (HR 1.38, 95% CI 1.11-1.72) and left colon (HR 0.60, 95% CI 0.48-0.75), and rectum (HR 0.59, 95% CI 0.42-0.82) with reference to right colon location but no association with other advanced features. **(D)** Histograms comparing frequency of a mutation in each of the top 10 mutated genes (panels) between index adenomas from right colon (blue), left colon (red), or rectum (green) showing significantly higher frequency of mutations in *KRAS* (padj<0.001), *TP53* (padj=0.013) in rectal adenomas compared to higher *BCOR* mutation frequency in right colon adenomas (padj=0.013) (Chi square test with Benjamini-Hochberg adjustment). **(E)** Inverse Kaplan Meier curves of time to detection of metachronous polyp or colorectal cancer (CRC) following polypectomy at index bowel screening colonoscopy showing no significant difference in time to event when index conventional adenomas were grouped by *KRAS* mutation status in right colon (p=0.486), left colon (p=0.499) or rectum (p=0.350).

When the frequency of the top 10 mutated genes in index adenomas was compared across right colon, left colon and rectal location (Figure 4D), there were significant differences in the frequency of *KRAS* mutation (42.9% vs. 27.5% vs. 50.0% respectively, padj<0.001), *TP53* mutation (7.6% vs. 16.2% vs. 25.6% respectively, padj=0.013) and *BCOR* mutation (12.6% vs. 4.6% vs. 9.0% respectively, padj=0.013). When *KRAS* mutation was stratified by index adenoma location (Figure 4E), there was no significant association with time to detection of future polyp or CRC between mutations status in the right colon (p=0.486), left colon (p=0.499), or rectum (p=0.350), with similar results for *TP53* mutation (Supplementary Figure 3) in the right colon (p=0.438), left colon (p=0.958) and rectum (p=0.435).

### Sequential accrual of classic adenoma-carcinoma sequence driver mutations in index conventional adenomas is associated with advanced histopathological features but not metachronous lesion risk

Given the association between *KRAS* mutation, *TP53* mutation and histologically advanced features in adenomas we then grouped lesions by perceived risk conferred by mutation. We considered a simplified schematic of the classic adenoma carcinoma sequence (Figure 5A) and grouped index adenomas into having no *APC* mutation (No-APC, n=69), an *APC* mutation alone (A, n=415), an *APC* and *KRAS* mutation (AK, n=180) and an *APC*, *KRAS* and *TP53* mutation (AKP, n=32)(Figure 5B). This method of ordinal grouping based on mutations in these 3 genes was further supported by a combination of the existing mutation frequency data in the index adenomas showing mutation frequency of 91% for *APC*, 33% for *KRAS* and 16% for *TP53*, and density plots of VAF estimates showing the highest median for *APC* at 0.24, with similar medians of 0.19 for *KRAS* and 0.18 for *TP53* (Supplementary Figure 4).

**Figure 5:**
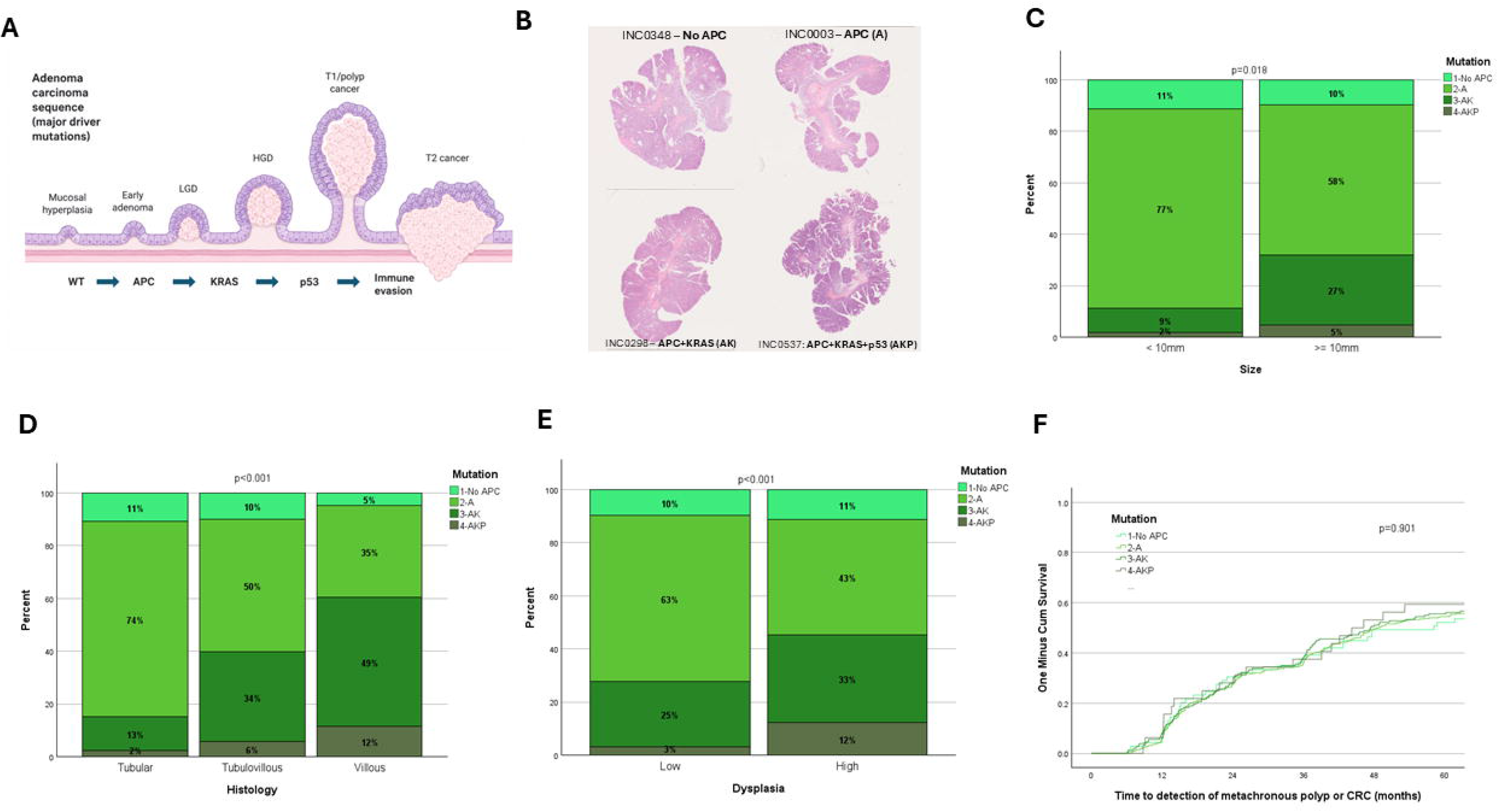
Accrual of mutations in classic colorectal carcinogenesis driver mutations is associated with advanced features but not future polyp or colorectal cancer in index conventional adenomas. **(A)** Simplified schematic of the conventional adenoma to carcinoma sequence from normal mucosa with a wild type (WT) landscape through accrual of driver mutations in *APC*, *KRAS* and *TP53* leading from early adenoma to increasing dysplasia and early stage (T1-T2) invasive colorectal cancer (CRC). **(B)** Haematoxylin and eosin stain (H&E) whole slide images of representative index conventional adenomas from the INCISE cohort with no *APC* mutation (No APC, n=69), *APC* mutation (A, n=415), *APC* and *KRAS* mutation (AK, n=180), *APC*, *KRAS* and *TP53* mutation (AKP, n=32). Stackplots of frequency of samples with no *APC* mutation (No APC), *APC* mutation (A), *APC* and *KRAS* mutation (AK), *APC*, *KRAS* and *TP53* mutation (AKP) show significantly higher proportions of accrued mutations in **(C)** samples ≥10mm in size (p=0.018), **(D)** tubulovillous and villous morphology (p<0.001), and **(E)** high grade dysplasia (p<0.001), unadjusted chi squared test. **(F)** Kaplan Meier curve of time to detection of metachronous polyp or colorectal cancer (CRC) following polypectomy at index bowel screening colonoscopy shows no significant difference in time to event when index conventional adenomas were grouped by accrued driver mutations (Log rank p=0.901).

Statistically significant stepwise increases in the proportion of index adenomas within each sequential mutation group was observed when compared to the three histological high-risk characteristics, size ≥10mm (Figure 5C, p=0.018), tubulovillous and villous features (Figure 5D, p<0.001) and HGD (Figure 5E, p<0.001). However, there was no association between the sequential mutation group and metachronous lesion risk during surveillance at time to event analysis (p=0.901). When index adenomas were grouped for every combination of these three driver mutations (Supplementary Figure 5) there was still no association with outcome (p=0.970).

## Discussion

This study, using one of the largest cohorts available for colorectal adenomas, reports that the accumulation of recognised driver mutations in conventional adenomas is strongly associated with advanced histopathological features but is not associated with the development of metachronous neoplasia during post-polypectomy surveillance. This central finding reinforces the interpretation that driver mutations primarily reflect lesion-level progression rather than the biological processes that govern future neoplasia risk. The most frequently mutated genes in this cohort aligned with previously characterised mismatch repair proficient (pMMR) conventional adenomas [31]. The associations between *KRAS* and *TP53* mutations and advanced histopathology are consistent with their canonical positioning in the adenoma-carcinoma sequence and with experimental evidence that sequential driver mutation accrual potentiates proliferative output, translational capacity, and stem cell expansion [32–33]. Critically, however, none of these mutation-defined features translated into an association with metachronous neoplasia risk, establishing a clear separation between determinants of lesion progression and determinants of future neoplastic susceptibility.

A further observation of this study is the presence of inter-lesional heterogeneity in TMB between index, synchronous, and metachronous adenomas and mutation frequency by colonic location. However, when clinicopathological variables were accounted for through propensity score matching at the polyp level, these differences were attenuated, with no significant difference observed between index and metachronous lesions. This suggests that much of the apparent quantitative mutational spatial and temporal heterogeneity is explained by differences in polyp size, histological grade, and architectural complexity, features that are themselves established correlates of mutational burden [20, 34]. Importantly, analyses using both per-patient and per-polyp frameworks showed no significant differences in driver mutation frequency between index, synchronous, and metachronous adenomas.

Our findings along with others can be reconciled within a unified model in which a colonic mucosal field [35], characterised by widespread low-frequency driver mutations [36], proteomic and epigenetic alterations [37–38] provides the background for the independent emergence of multiple adenomas. Within this field, individual lesions arise as distinct clonal expansions, but subsequently evolve under spatially and temporally variable selective pressures [39]. Resulting inter- and intra-lesional heterogeneity is anticipated by the polyclonal origins of intestinal adenomas [18], sub-clonal competition within individual colorectal adenomas [14], and the different cellular and immune microenvironmental compositions observed between adenomas [15, 40]. However, the relative conservation of key driver mutations in adenomas across space and time suggests that, despite this heterogeneity, tumour evolution remains constrained to a limited set of oncogenic pathways, such as activated *Wnt* signalling, reflecting common biological requirements for adenoma formation and progression [16].

Taken together this has important implications for biomarker development in post-polypectomy surveillance. Histopathological features of the index adenoma remain the most actionable predictors of metachronous lesion detection [41]. However, the British Society of Gastroenterology (BSG) 2020 guidelines [42] were not predictive of metachronous lesions detected beyond two years from the index colonoscopy in the INCISE cohort [9], underscoring the need for additional biomarkers capable of capturing field-wide susceptibility rather than lesion-specific mutational state. The stability of driver mutation frequency across lesions indicates that targeted mutation profiling alone is unlikely to distinguish patients at risk of future neoplasia. Approaches with greater potential could include integrating gene expression signatures, genomic-methylation data [21, 37], and immune microenvironmental characterisation of index resected adenoma and/or surrounding mucosal tissue [43]. The integration of such data with host demographic and exposure information may ultimately yield composite risk models with sufficient discriminative ability to personalise surveillance intervals.

This study has a number of limitations. The use of a cancer-specific targeted gene panel introduces systematic ascertainment bias, enriching for point mutations in canonical driver genes whilst omitting structural variants, copy-number alterations, and epigenomic features. The whole lesion “bulk” nature of the mutation analysis will likely average mutation frequency and pre-malignant biologies of different regions of adenoma epithelium and lamina propria preventing meaningful assessment of intra-lesional heterogeneity. The driver annotation used means that detection of novel drivers in this setting is unlikely but was not the focus of this analysis. The calculation of TMB from a targeted panel is subject to sampling bias, however the exclusion of drivers should reduce the impact of positive selection and hotspot clustering, with a 3.96Mb panel size adequate for such an estimate [44]. The lack of copy number and cellularity data mean that the calculated VAF for *APC*, *KRAS* and *TP53* is an estimate and may be subject to error [45]. Very few of the included samples were <10mm in size, as larger lesions were perceived to be more likely to undergo successful DNA extraction. This may however come to mimic evolving clinical pathways in which adenomas 5mm or smaller will be discarded following polypectomy without the need for histopathological review [46]. The relatively small number of sampled metachronous adenomas reflects the protracted timescale of surveillance programmes and the practical constraints of tissue availability from diminutive surveillance polypectomy specimens, limiting statistical power to detect modest effect sizes and introducing the possibility of type II error for subgroup analyses. The propensity score matching analysis, whilst addressing confounding by lesion-level clinicopathological features, cannot account for unmeasured variables that may independently influence both mutational burden and metachronous risk.

In conclusion, this study confirms that classical driver mutations are robust indicators of adenoma progression, while demonstrating their limited value in predicting metachronous lesion risk. Despite observable inter-lesional heterogeneity in clinicopathological features between lesions, there exists a remarkably conserved driver mutation landscape across index, synchronous, and metachronous adenomas. Collectively, these findings underscore a critical limitation of mutation only profiling and provide a strong rationale for integrated, multi- dimensional risk stratification approaches to more accurately inform personalised post-polypectomy surveillance and improve patient outcomes.

## Data availability statement

Anonymised clinical and pathological data are available on the secure Glasgow Safe Haven TRE platform. Access can be arranged by application to the authors and via the Glasgow SafeHaven TRE following ethical approval and the completion of mandatory information governance modules. Mutational data are available publicly at (https://www.ebi.ac.uk/biostudies/ArrayExpress/studies/E-MTAB-15346?query=E-MTAB-15346, accession number E-MTAB-15346) and IHC data access can be discussed on reasonable application to the corresponding author.

## Ethics

Ethical approval was obtained for data (GSH/20/CO/002) and tissue analysis (22/WS/0020).

## Acknowledgements

The authors and the **IN**tegrated **T**e**C**hnologies for **I**mproved Polyp **S**urveillanc**E** (INCISE) collaborative group thank Glasgow Tissue Research Facility, specifically Dr Pamela McCall, Dr Jennifer Hay, Mr Jakub Jawny, Dr Hannah Morgan and Mr Scott Murray. We thank Greater Glasgow and Clyde Biorepository including Ms Clare Orange for help in identifying and retrieving archival pathology tissue. We thank Mr William Sloan of NHS Greater Glasgow and Clyde for help in patient identification. We thank the patients for use of their tissue.

## Contributors

STM, GL and JE conceived and designed the study. STM, LSS, AA and SSFA analysed and interpreted the results. STM supervised the statistical analysis. NM provided expert pathological input and specimen review. STM wrote the manuscript. LSS, AA, SSFA, EP, PD, NM, MJ, GL, and JE critically reviewed the manuscript. STM, GL and JE supervised the study. GL provided administrative and project management support. ECP managed the PPIE aspects. All authors critically read and approved the paper. STM is the guarantor of the manuscript.

## Funding

Innovate UK: Awards 10054829 and 105858 Guts UK: ECR2023_03

Medical Research Scotland: PhD Studentship - PhD-50246-2020 Chief Scientist Office: Early Postdoctoral Fellowship - EPD/25/14 Cancer Research UK Scotland Institute: CTRQQR-2021\100006

### Beatson Cancer Charity 330619-01/ 25-26-076

CRUK Early Detection Programme and Bowel Babe Fund (EDDPGM-Nov25/100003).

## Patient and public involvement

The results of this study were presented to the Glasgow Colorectal PPIE group (https://www.gla.ac.uk/research/az/incise/forpatientsandthepublic/). The authors thank the group members for their contribution to the wording and emphasis of the reporting and their ongoing help with dissemination.

