## Supplementary data for "Classical driver mutations are not associated with metachronous lesion risk in patients undergoing post-polypectomy surveillance following removal of conventional adenomas in a bowel screening setting"

**A**

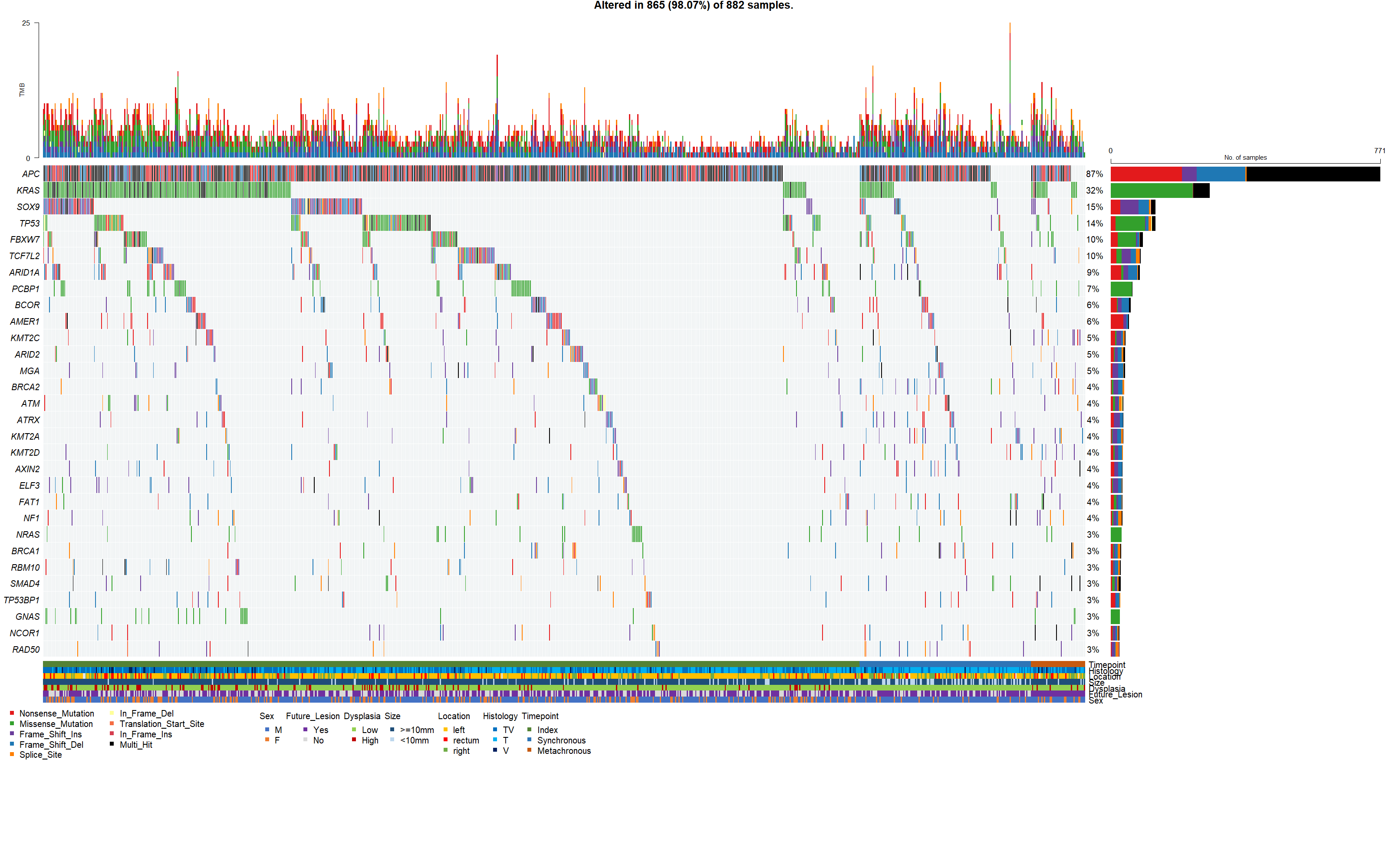

**B**

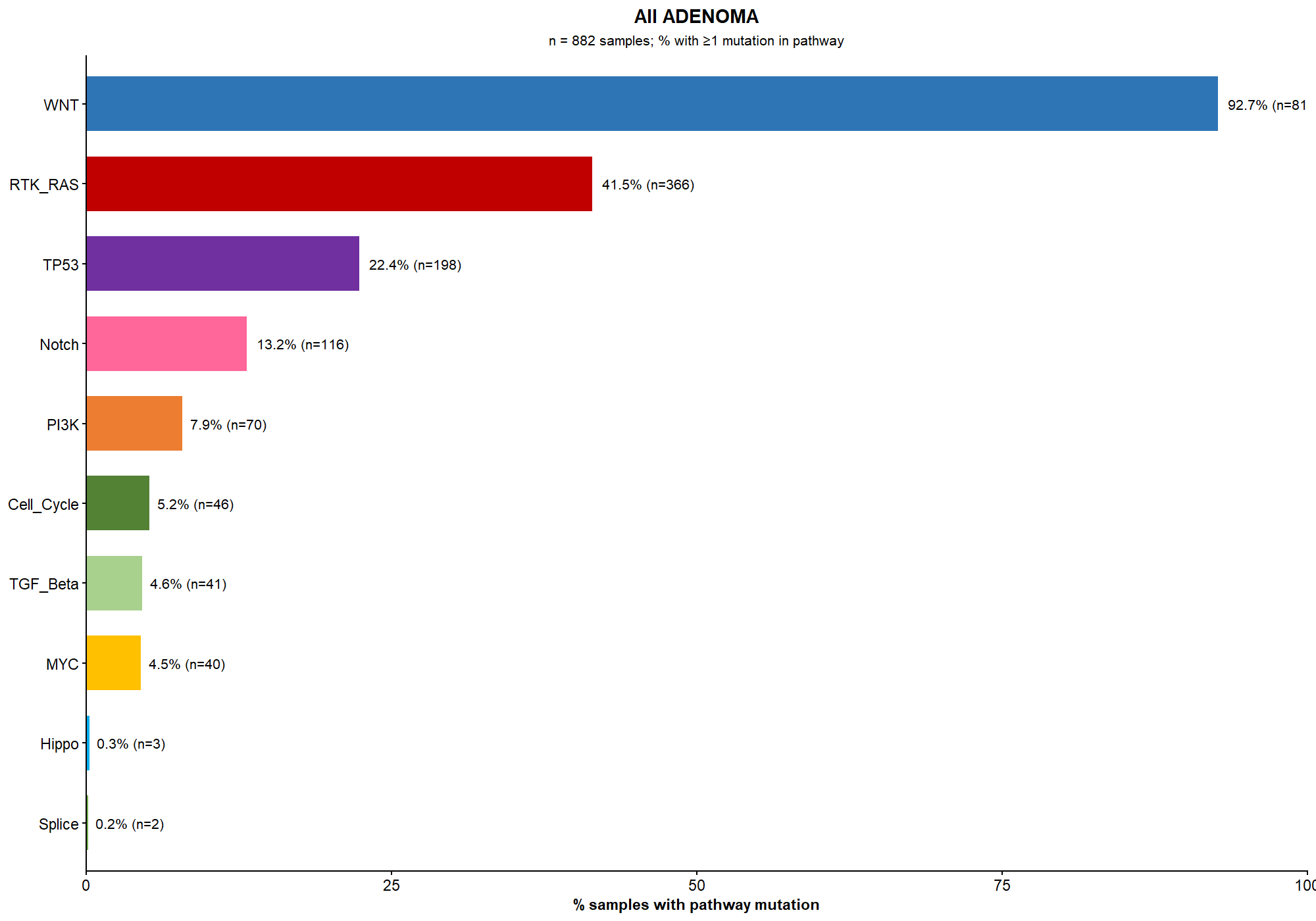

Supplementary Figure 1: (A) Oncoplot of top 30 mutated genes in conventional adenoma samples (n=895). (B) Oncogenic pathway analysis histogram displaying the proportion of samples harbouring at least one mutation in curated gene sets (Sanchez-Vega et al. 2018).

**A**

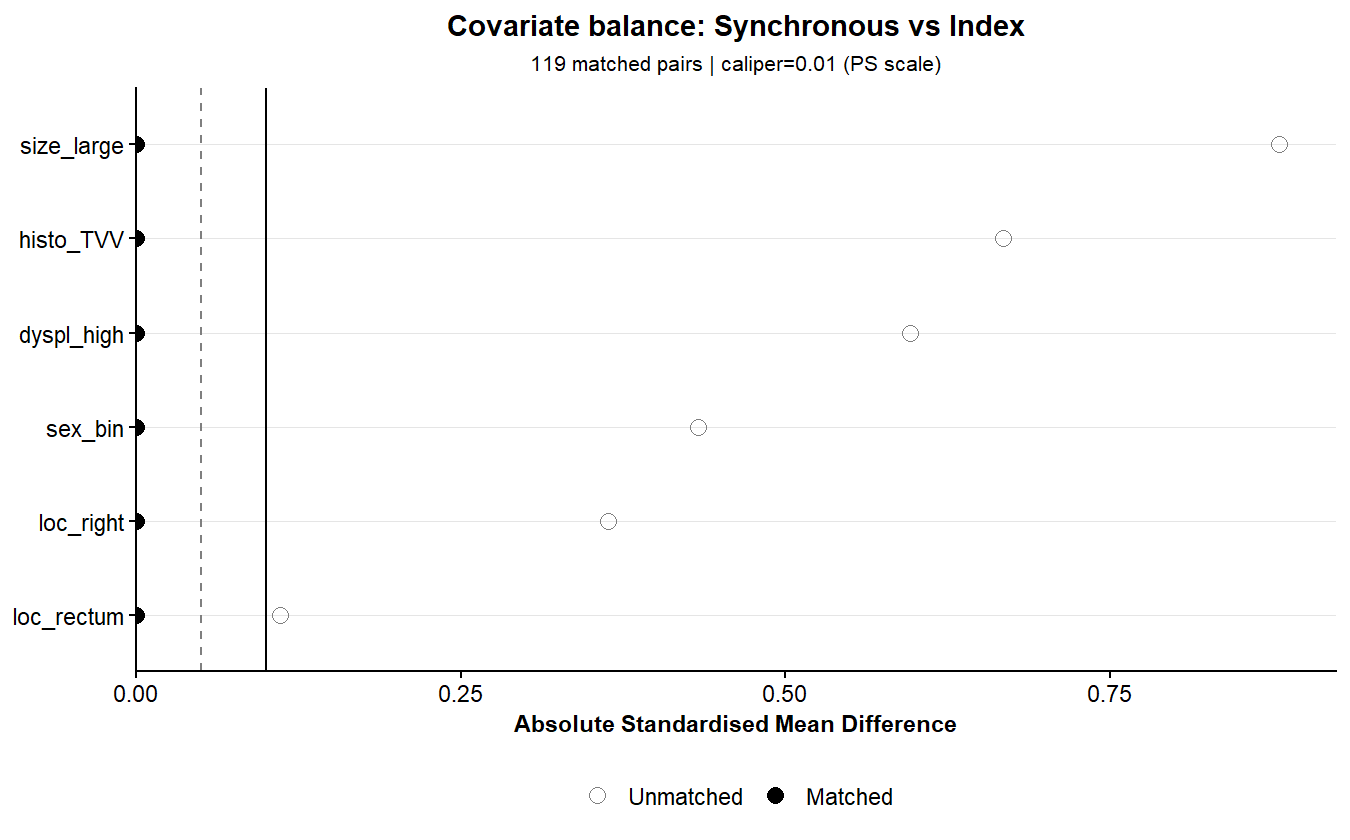

**B**

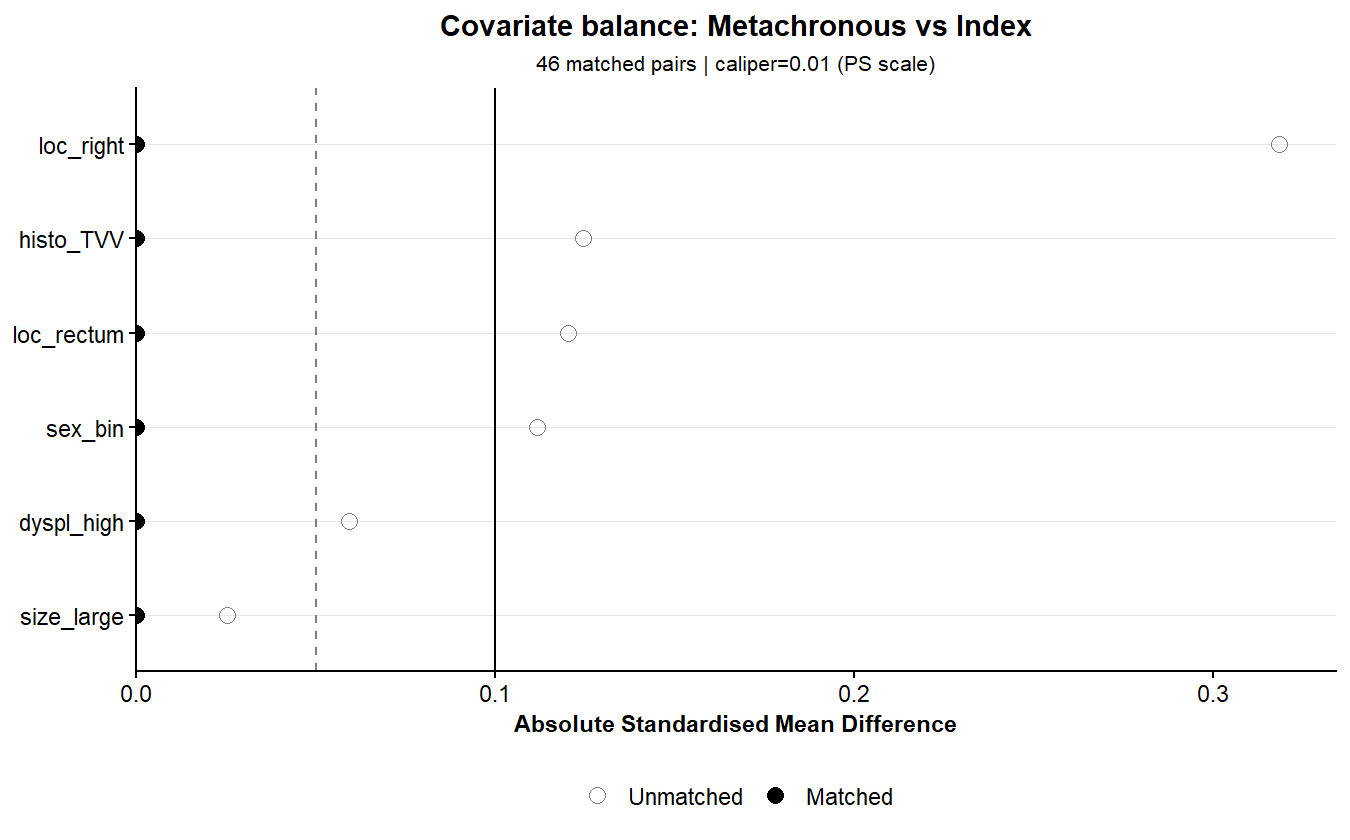

Supplementary Figure 2: Covariate balance charts showing reduction in Standardised Mean Difference in propensity score (x-axis) between (A) index and synchronous and (B) index and metachronous adenoma samples before (empty dots) and after (filled dots) one to one propensity score matching per polyp using an absolute caliper of 0.01, without replacement, for each given variable used to generate the propensity score (y axis).

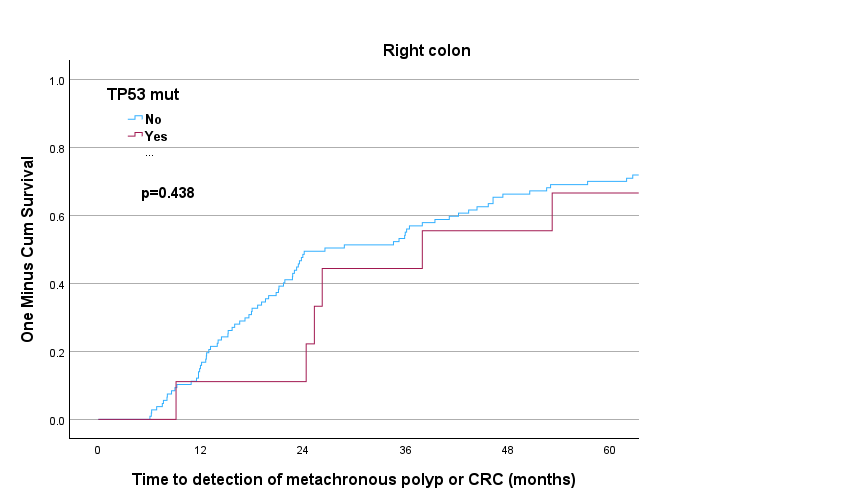

**A**

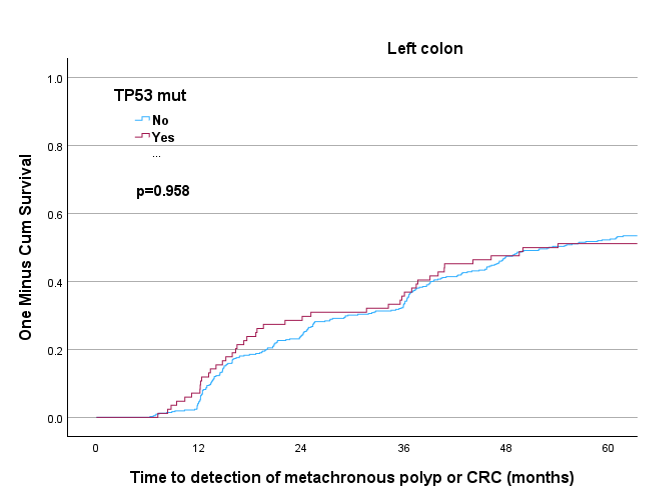

**B**

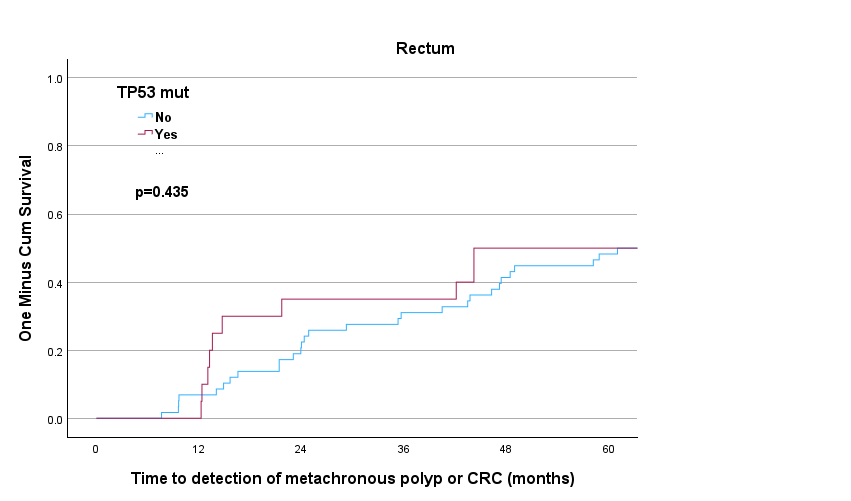

**C**

Supplementary Figure 3: Kaplan Meier curve showing no significant difference in time to detection of metachronous polyp or colorectal cancer (CRC) when comparing index adenomas with and without TP53 mutations in the (A) right colon (p=0.438), (B) left colon (p=0.958) or (C) rectum (p=0.435).

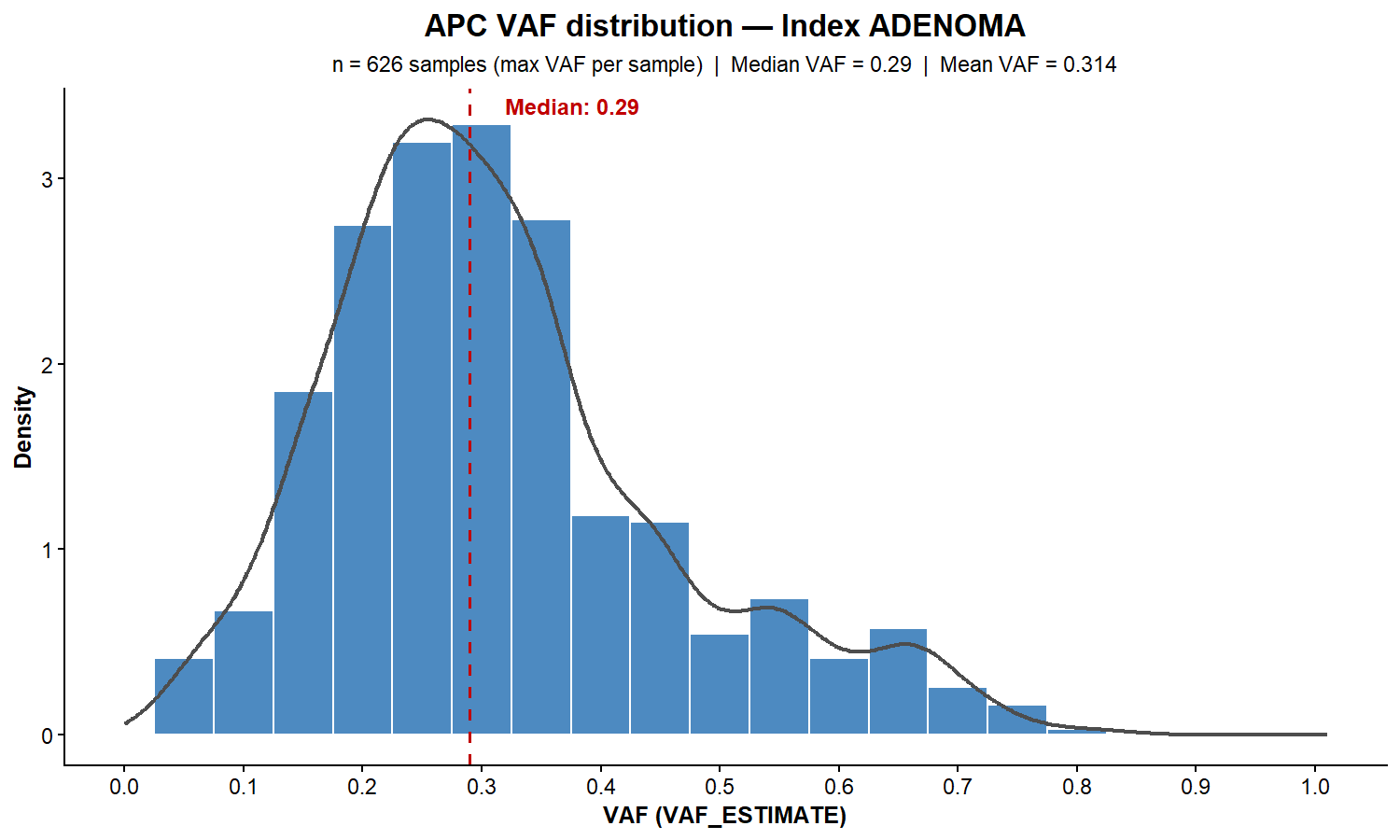
**A**

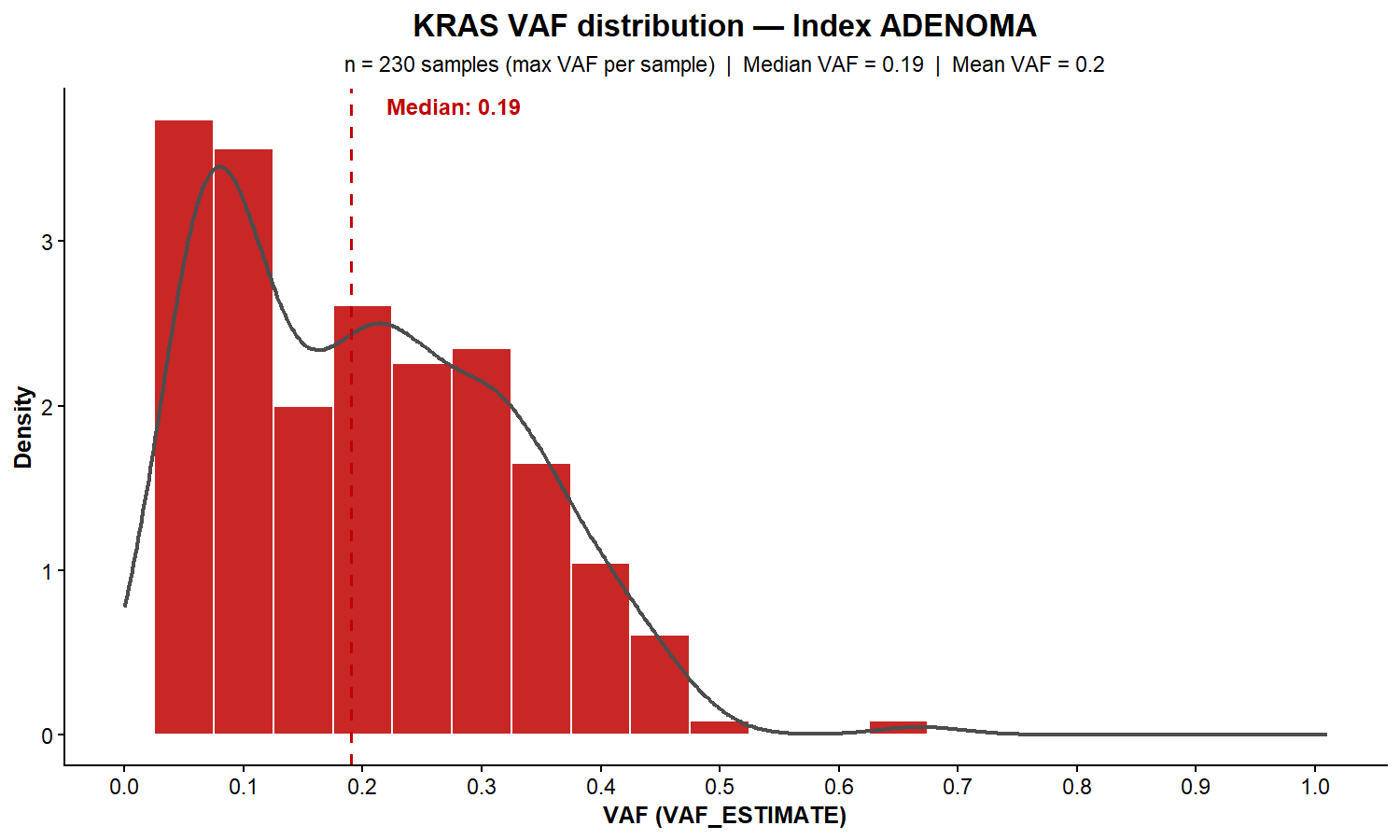
**B**

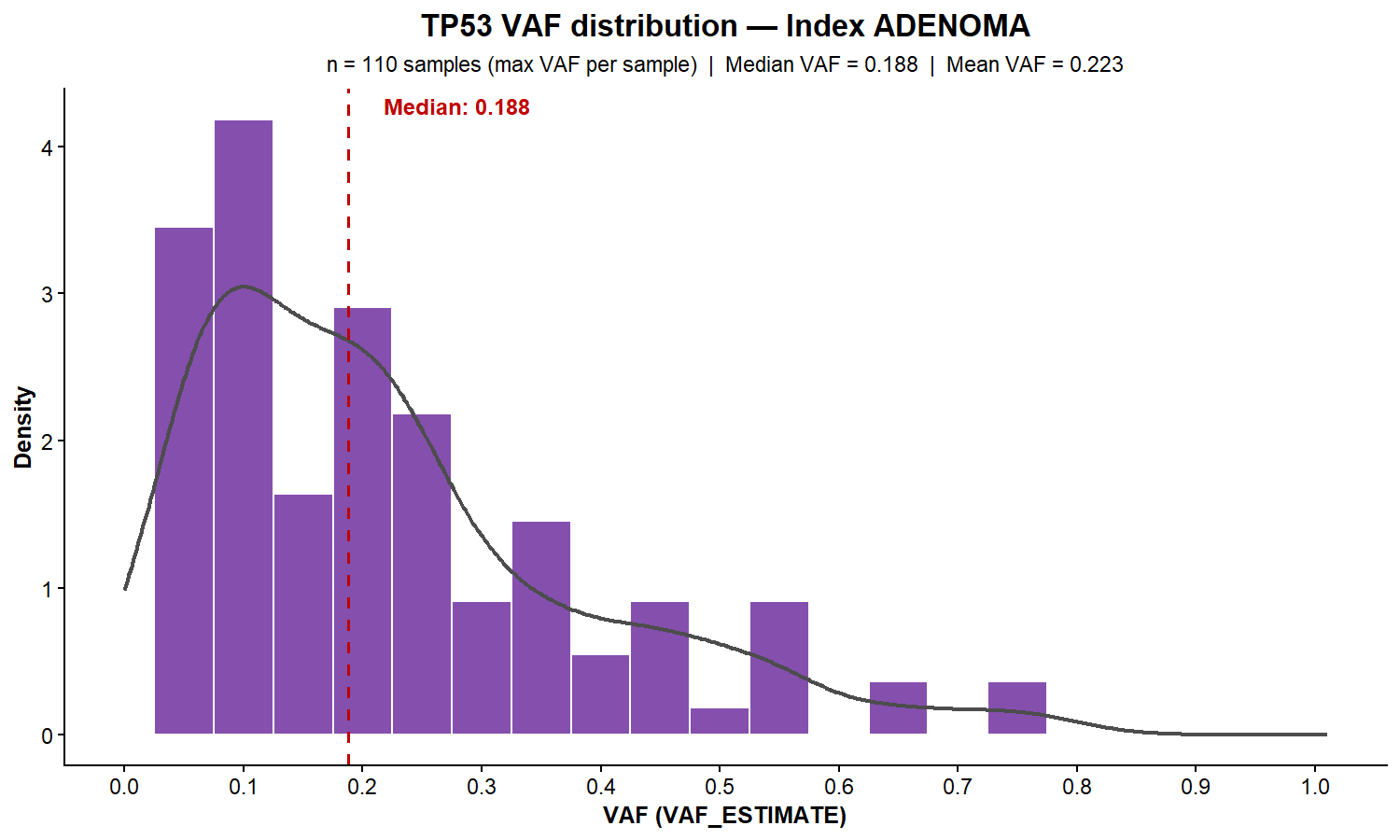
**C**

Supplementary Figure 4: Estimated variant allele frequency (VAF) density plots for **(A)** A*PC*, **(B)** *KRAS*, and **(C)** *TP53* mutations in index conventional adenomas

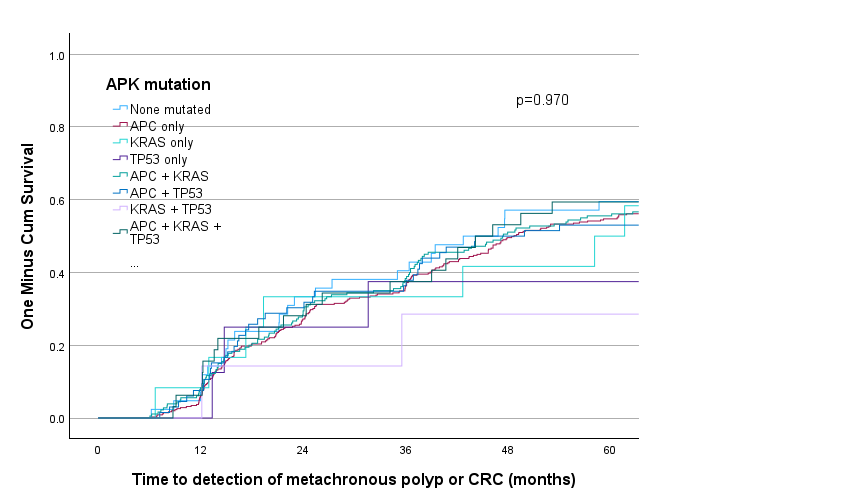

Supplementary Figure 5: Kaplan Meier curve showing no significant difference (log rank p=0.970) in time to detection of metachronous polyp or colorectal cancer (CRC) when comparing index adenomas grouped by mutation in *APC*, *KRAS* or *TP53*.

| KRAS mutation type | Right colon | Left colon | Rectum | p | p adj |
| --- | --- | --- | --- | --- | --- |
| N samples | 119 | 501 | 78 |  |  |
| G12A | 4 (3.4%) | 9 (1.8%) | 2 (2.6%) | 0.442 | 0.68 |
| G12C | 3 (2.5%) | 9 (1.8%) | 2 (2.6%) | 0.701 | 0.779 |
| G12D | 8 (6.7%) | 37 (7.4%) | 8 (10.3%) | 0.623 | 0.737 |
| G12S | 2 (1.7%) | 4 (0.8%) | 3 (3.8%) | 0.044 | 0.18 |
| G12V | 16 (13.4%) | 39 (7.8%) | 12 (15.4%) | 0.031 | 0.18 |
| Q61H | 0 (0%) | 8 (1.6%) | 2 (2.6%) | 0.252 | 0.513 |
| A146P | 1 (0.8%) | 1 (0.2%) | 0 (0%) | 0.487 | 0.696 |
| A146T | 3 (2.5%) | 2 (0.4%) | 2 (2.6%) | 0.028 | 0.18 |
| A146V | 2 (1.7%) | 0 (0%) | 0 (0%) | 0.045 | 0.18 |
| G12R | 1 (0.8%) | 2 (0.4%) | 0 (0%) | 0.627 | 0.737 |
| G13D | 7 (5.9%) | 22 (4.4%) | 6 (7.7%) | 0.412 | 0.68 |
| G13R | 0 (0%) | 0 (0%) | 1 (1.3%) | 0.125 | 0.416 |
| G60D | 0 (0%) | 1 (0.2%) | 1 (1.3%) | 0.237 | 0.513 |
| K117N | 1 (0.8%) | 2 (0.4%) | 0 (0%) | 0.617 | 0.737 |
| K117R | 1 (0.8%) | 0 (0%) | 0 (0%) | 0.264 | 0.513 |
| K5E | 0 (0%) | 1 (0.2%) | 0 (0%) | 1 | 1 |
| L19F | 1 (0.8%) | 0 (0%) | 0 (0%) | 0.28 | 0.513 |
| Q61R | 1 (0.8%) | 0 (0%) | 0 (0%) | 0.282 | 0.513 |
| R68S | 0 (0%) | 1 (0.2%) | 0 (0%) | 1 | 1 |
| No mutation | 68 (57.1%) | 363 (72.5%) | 39 (50%) | <0.001 | <0.001 |

Supplementary table 1: Comparison of proportion of frequency of specific KRAS mutations defined by protein change in index adenomas, compared across right colon, left colon and rectal location (chi square test with Benjamini Hochberg adjustment).
